# Perceived Social Support and Self-Efficacy as Mediators Between Health Literacy and Quality of Life Among Middle-Aged and Older Adults with Hypertension: A Cross-Sectional Study in Six Central Provinces of China

**DOI:** 10.64898/2026.06.06.26355051

**Authors:** Yue Zhao, Yun Yun, Tingting Bai, Luyao Xiong, Yanan Ruan, Hui Zhao, Weixian Wang, FuZhi Wang

## Abstract

**Objective:** The onset of hypertension occurs at a younger age in China, and the relationship between health literacy and quality of life among middle-aged and older hypertensive patients remains unclear. This study explored whether perceived social support and self-efficacy mediate the association between health literacy and quality of life in middle-aged and older hypertensive patients.

**Methods:** A questionnaire was administered to 1,015 middle-aged and older hypertensive adults from communities in six central provinces of China. The EQ-5D scale, Perceived Social Support (PSS) scale, Self-Efficacy Scale (SES), and Health Literacy Scale (HLS) were used to assess quality of life, social support, self-efficacy, and health literacy, respectively. Mplus 8.3 software was used to construct a structural equation model for path analysis.

**Results:** The mean PSS, SES, HLS, EQ-5D, and EQ-VAS scores were 15.57±3.45, 10.61±2.41, 9.49±2.86, 0.88±0.18, and 71.06±17.49, respectively. Health literacy and quality of life scores significantly differed among middle-aged and older hypertensive patients, and both showed positive correlations with perceived social support and self-efficacy (both P<0.001). Perceived social support and self-efficacy exhibited a chain mediated effect on the relationship between health literacy and quality of life (EQ-5D utility index and EQ-VAS), accounting for 28.57% of the total effect of the EQ-5D utility index and 27.26% of that of the EQ-VAS. This study is the first to elucidate the mechanism by which health literacy influences quality of life in middle-aged and older hypertensive patients through the chain-mediated effect of perceived social support and self-efficacy.

**Conclusion:** Health literacy is significantly correlated with quality of life in middle-aged and older hypertensive patients. This correlation can directly or indirectly explain the impact on quality of life through mediating pathways involving perceived social support and self-efficacy.

## Background

With the continuous aging of the population, chronic diseases among older individuals have become an increasingly prominent global issue. As individuals enter the latent stage of aging during middle age, health concerns among this demographic also warrant heightened attention. According to the World Health Organization’s first global hypertension report ^[1]^, hypertension, a common chronic disease, affects over 1 billion people worldwide. Data show that the number of hypertensive patients in China has reached 245 million, with the overall prevalence rate continuing to rise ^[2]^. Hypertension is closely associated with multi-organ and tissue damage and significantly increases the risk of developing diabetes, chronic kidney disease, and other cardiovascular and cerebrovascular diseases. Thus, hypertension has emerged as a major public health threat ^[3]^; it not only correlates with increased physical suffering and treatment burden for patients but also contributes to diminished quality of life.

The effective management of hypertension not only relies on pharmacological treatment but also is closely associated with patients’ health literacy. Health literacy refers to an individual’s ability to acquire, understand, and utilize health information or services to make decisions that improve and maintain their health behaviours ^[4]^. Existing studies indicate a significant positive correlation between the level of health literacy in hypertensive patients and the efficacy of blood pressure management ^[5]^. Improvements in patients’ quality of life are also influenced by external interventions and self-efficacy ^[6]^. Self-efficacy denotes an individual’s confidence in their ability to achieve specific goals ^[7]^. Research demonstrates that self-efficacy is positively correlated with disease management in hypertensive patients and associated with a reduced risk of complications ^[8]^. In addition to self-efficacy, perceived social support, referring to the support individuals receive from family, friends, and others, serves as a critical psychosocial factor ^[9]^. Relevant studies have also confirmed a significant positive correlation between perceived social support and quality of life in hypertensive patients ^[10]^. These findings suggest that enhancing middle-aged and older individuals’ perception of social support may contribute to improved quality of life in hypertensive patients while potentially alleviating the caregiving burden on families and society.

The interactions among health literacy, self-efficacy, and perceived social support have also garnered widespread attention. Studies indicate that the association between health literacy and quality of life may be mediated by social support ^[11]^. Further research confirms that patients who receive higher levels of social support tend to exhibit better quality of life (a positive correlation exists between these two factors) ^[12]^. Additionally, studies demonstrate that self-efficacy mediates the relationship between health literacy and quality of life ^[13]^. Furthermore, research shows that the perceived social support and self-efficacy levels among patients mutually reinforce one another. The synergistic interaction between these two factors is significantly correlated with improved quality of life, suggesting that enhancing both factors may create a positive feedback loop closely linked to increased well-being ^[14]^. However, these studies still have specific limitations. First, most studies focus solely on the analysis of individual mediating pathways, failing to integrate health literacy, perceived social support, self-efficacy, and quality of life into a unified research framework. Consequently, the cascading mechanisms underlying their interactions remain unclear. Second, recent studies on middle-aged and older hypertensive patients have only validated the independent effects of each variable. No integrated cascading mediation model encompassing all four variables based on social cognitive theory has been established, making it difficult to elucidate the complete pathway through which health literacy influences quality of life in this population.

This study investigated middle-aged and older hypertensive patients. We constructed a structural equation model based on social cognitive theory to examine the mediating effects of perceived social support and self-efficacy on the relationship between patients’ health literacy and quality of life. This study provides theoretical support for decision-making to develop targeted interventions to improve quality of life of middle-aged and older hypertensive patients.

This study reports the construction of an analytical framework based on Bandura’s social cognitive theory, with the core objective of elucidating the dynamic interactions among individual factors, environmental factors, and health behaviours, thereby providing a fundamental theoretical basis for model development ^[15]^. The theory posits that environmental factors encompass the provision of external resources such as perceived social support, while individual factors include cognitive and ability dimensions, such as health literacy and self-efficacy. As key psychological mediators in this theory, self-efficacy and perceived social support serve as bridges that link individual cognitive characteristics to health outcomes.

On the basis of this theoretical framework, this study further elucidates a chain-mediated pathway comprising health literacy → perceived social support → self-efficacy → quality of life. Both robust theoretical validation of and empirical support for this pathway are presented. From a theoretical perspective, the “triple interaction theory” in social cognition emphasizes the pivotal role of external environmental factors in shaping and developing individual psychological capacities. This directly validates the scientific rationale for positioning perceived social support (an environmental factor) as an antecedent variable of self-efficacy (an individual psychological factor), rather than viewing them as parallel or inverse relationships.

Empirical studies have provided robust support for each link in this chain. This logic has been empirically validated in hypertensive populations. Research findings have revealed a significant positive correlation between perceived social support (an environmental factor) and self-efficacy (an individual psychological factor), with self-efficacy serving as a mediating factor between social support and self-management ^[16]^. This provides crucial evidence for the view that “perceived social support is a precursor to self-efficacy.” Furthermore, each link in this chain has been empirically supported. Wang et al. ^[17]^ conducted a study involving 401 community hypertensive patients and found a significant positive correlation between health literacy and social support, confirming that health literacy enhances patients’ ability to perceive social support. Through structural equation modelling of rural hypertensive patients in China, Zhang et al. ^[18]^ demonstrated that health literacy, social support, and self-efficacy collectively exert a positive impact on quality of life via this chain, directly supporting the construction of a comprehensive chain structure in this study.

Overall, this study adopts social cognitive theory as its core guiding framework. Via the integration of existing empirical research, a chain-mediated model that not only fully aligns with this theoretical core but also effectively addresses the limitations of previous studies in explaining the synergistic pathways among explanatory variables has been constructed. This methodological approach establishes a solid theoretical foundation while demonstrating rigorous scientific design.

### Materials and Methods Data sources

The research data were derived from the 2024 China Resident Psychology and Behavior Survey (PBICR) ^[19]^. This survey aimed to establish a database through a large-scale, multicentre, reproducible, and nationwide cross-sectional study, providing reliable data support for research across various fields and enabling a comprehensive and systematic understanding of the public’s physical and mental health status. This study has been approved by the Shanghai Jiao Tong University Ethics Review Committee (H20240237I).This survey was conducted from June 20,2024, to August 31,2024. Data were collected via anonymous questionnaires, with no personal identification information obtained. Data management and usage strictly complied with the *Statistics Law of the People’s Republic of China*^[20]^, and all information was kept strictly confidential. The research data will only be made available for open access and sharing after the complete anonymization of personal information and are restricted to academic research purposes only. The researchers retrieved the data for research purposes on October 1, 2025. No personal information will be disclosed upon publication of the research findings, and participants will not suffer any adverse effects as a result. After completing the project team survey and related tasks, participants may apply for data access. Upon approval by the project team, participants must sign a data confidentiality agreement. Once the agreement takes effect, the project leader will directly send the research data to the applicant via email, ensuring secure and controlled data access.

### Research Subjects

#### 1. Inclusion Criteria and Exclusion Criteria

The inclusion criteria for this study sample were as follows: (1) age 45 years or older^[21]^; (2) Chinese nationality; (3) permanent residence in China (annual time spent outside China ≤ 1 month); (4) voluntary participation in the study and completion of an informed consent form; (5) ability to complete the online questionnaire independently or with investigator assistance; (6) understanding of the meaning expressed in each questionnaire item; and (7) diagnosis of hypertension. The exclusion criteria for participants were as follows: (1) individuals with impaired consciousness or mental disorders; (2) individuals with cognitive impairment; (3) individuals currently participating in other similar research studies; and (4) individuals unwilling to cooperate.

This survey questionnaire was completed by 11,944 participants from 82 cities across six provinces in central China: Shanxi, Anhui, Jiangxi, Henan, Hubei, and Hunan. On the basis of the inclusion and exclusion criteria, this study ultimately included data from 1,015 middle-aged and older hypertensive patients.

#### 2. Sampling Method

This study was based on the Psychology and Behavior Investigation of Chinese Residents (PBICR), and a multistage stratified sampling method was used for the survey. First, in six provinces of central China, the number of cities to be sampled was determined on the basis of each province’s population base. The random number table method was used to select a total of 80 cities. Afterwards, within these 80 sampled cities, the number of communities to be sampled was determined on the basis of the population base of the first-level administrative region in which the city is located. In each city sampled from the provinces, 228 communities were selected in total, with a 3:2 urban community: rural community ratio. The final recovered questionnaires were screened on the basis of the criteria of “age ≥45 years and diagnosed hypertension.”

#### 3. Sample Size Calculation

This study employed a chain mediation analysis method based on structural equation modelling. According to previously described methodology ^[22]^, the sample size was estimated using a publicly available calculator. The core model parameters included 16 observed variables, 4 latent variables, a common mediation effect size of 0.15 in the mediation model, α = 0.05, and a power of 0.8. The calculator yielded a minimum sample size of 438 participants. Ultimately, the study enrolled 1,015 subjects, achieving 2.3 times the minimum sample size requirement. This adequate sample size ensured the reliability of structural equation model parameter estimation and the stability of model fitting, thereby meeting the analytical demands of the study.

#### 4. Standardized Quality Control for Blood Pressure Measurement

In this study, blood pressure was measured and questionnaires were administered at community sites. To ensure the accuracy and consistency of the measurement results, standardized quality control measures were implemented: All measurements were scheduled between 9:00 AM and 11:30 AM daily. Subjects were required to sit still for 30 minutes upon arrival, and smoking, vigorous exercise, and the consumption of caffeine-containing beverages were prohibited prior to measurement. Measurements were performed using the right upper arm, with the arm kept at heart level to ensure proper cuff coverage over the brachial artery. Two readings were recorded for each measurement (with an interval of 1–2 minutes); if the difference in blood pressure exceeded 5 mmHg, an additional reading was recorded. The average of the last two readings constituted the valid data. All electronic blood pressure monitors were pre-calibrated. Measurement personnel received standardized training prior to data collection. On-site supervisors monitored the procedures in real time and verified the data to strictly control measurement errors.

### Research Tools

All the scales used in this study are survey instruments employed within the framework of the Chinese Residents’ Mental Health and Behavioral Survey Project. They are tailored to the cognitive characteristics and questionnaire completion burden of middle-aged and older hypertensive individuals. The short-form versions of these scales significantly increase response rates and completion quality. Furthermore, all short-form scales utilized in this study have undergone rigorous validation, demonstrating excellent consistency in reliability and validity with their full-length counterparts, thereby effectively ensuring measurement accuracy. Upon verification, these survey tools have no authorization issues. All short-form versions of the scales consist of positive items only, with no negative items. Among these, the EuroqQol 5 dimension 5 level (EQ-5D-5L) was the official scale used, and the utility index was directly calculated using the formula. No additional reverse scoring was needed during data analysis.

#### 1. General Sociodemographic Information

The demographic indicators extracted from the 2024 Psychology and Behavior Investigation of Chinese Residents data include 12 items: gender, age, educational attainment, marital status, occupation, household registration status, monthly income, smoking status, alcohol consumption status, added salt in food, hypertension classification, and hypertension complications.

#### 2. Perceived Social Support

Perceived social support was measured using the 3-item short form of the Multidimensional Scale of Perceived Social Support (MSPSS). The MSPSS was developed by Zimet et al. ^[23]^ and simplified and revised by Wu et al. ^[24]^. It is primarily used to assess individuals’ perceived levels of support from three sources: family, friends, and others. Items were scored on a 7-point Likert scale ranging from 1 (strongly disagree) to 7 (strongly agree). The total score ranged from 3 to 21 points, with higher scores indicating stronger perceived social support. This study yielded a Cronbach α coefficient of 0.867.

#### 3. Self-Efficacy

Self-efficacy was measured using the 3-item New General Self-Efficacy Scale-Short Form (NGSES-SF). The NGSES-SF was developed by Chen et al. ^[25]^ and subsequently revised for the Chinese population by Feng et al. ^[26]^; this scale is mainly used to assess an individual’s confidence in their ability to successfully complete tasks or achieve goals in specific situations. Responses to each item were assessed on a 5-point Likert scale ranging from 1 (strongly disagree) to 5 (strongly agree). The total score ranged from 3 to 15 points, with higher scores indicating better overall performance. The Cronbach alpha coefficient measured in this study was 0.894.

#### 4. Health Literacy

Health literacy was assessed using the four-item version of the Health Literacy Scale-Short Form (HLS-SF4). This scale was developed by Sun et al. ^[27]^ and subsequently simplified and revised by Sun Xiaonan et al. ^[28]^. The HLS-SF4 scale is primarily used to evaluate respondents’ abilities to acquire, process, understand, and apply health-related information. Responses to each item were scored using a 4-point Likert scale, ranging from 1 (very difficult) to 4 (very easy). The total score ranged from 0 to 12, with higher scores indicating greater proficiency in health knowledge dissemination. The Cronbach’s α coefficient obtained in this study was 0.876.

#### 5. Quality of Life

Quality of life was measured using the EuroQol five-dimensional five-level questionnaire (EQ-5D-5L), which comprises two components: the EQ-5D descriptive system and the EuroQol Visual Analog Scale (EQ-VAS) ^[29]^. The EQ-5D descriptive system comprises five dimensions: mobility, self-care, usual activities, pain/discomfort, and anxiety/depression. The graded results across dimensions are integrated and converted into a single utility index to comprehensively reflect the health status of the study subjects. The utility index conversion system adopted in this study is based on the valuation standards for urban populations in mainland China, with a value range of -0.391 to 1.000. Here, 1.000 represents the best possible quality of life, 0 indicates death, and values <0 denote health states worse than death ^[30]^. Additionally, the EQ-VAS provides a quantitative description of respondents’ overall health perceptions, allowing patients to rate their health status on a scale from 0 (the worst health state imaginable) to 100 (the best health state imaginable). Higher EQ-5D utility index and EQ-VAS scores indicate better quality of life for survey participants. The Cronbach α coefficient for the EQ-5D-5L was 0.819.

### Statistical Analysis

Statistical analysis was conducted using SPSS 26.0 software. Categorical data are presented as frequencies and percentages, while continuous data are expressed as (X ± s). Data analysis was carried out via Pearson correlation analysis. Structural equation modelling was performed using Mplus 8.3 software for path analysis, with parameter estimation conducted via maximum likelihood estimation and mediation testing conducted using the bootstrap method. The structural equation model constructed in this study did not include demographic or disease-related variables as covariates; it comprised only four core latent variables—health literacy (an independent variable), perceived social support (a mediating variable), self-efficacy (a mediating variable), and quality of life (a dependent variable)—and analysed their path relationships. The primary objective of this study was to validate the relevant mediating mechanisms.

## Results

### Results of the descriptive statistical analysis

Regarding basic demographic data, individuals aged 65 and above accounted for a substantial proportion of participants at 48.08% (488/1015); males constituted 57.24% of the participants (581/1015), exceeding the proportion of females and individuals with other demographic characteristics. Primary school education was the predominant educational level, accounting for 25.52% of participants (259/1015). A total of 87.29% (886/1015) of participants were married; other occupations accounted for a relatively high proportion of participants at 20.10% (204/1015). Participants with monthly earnings ≤3,000 yuan and 3,001–4,000 yuan accounted for 81.67% of the participants (551/1015), significantly higher than the proportion of individuals in other income brackets. Non-agricultural household registration accounted for 53.99% of participants (548/1015), while agricultural household registration accounted for 46.01% (467/1015). In terms of lifestyle and habits, both smoking cessation and alcohol abstinence rates were high, at 74.39% (755/1015) and 72.22% (733/1015), respectively. Additionally, 58.23% (591/1015) of middle-aged and older individuals never or rarely added extra salt to their diet. Regarding disease-related conditions, 69.75% (708/1015) of middle-aged and older individuals had stage 1 hypertension; moreover, 49.36% (501/1015) of participants did not exhibit hypertension-related complications (see Table 1).

**Table 1.**
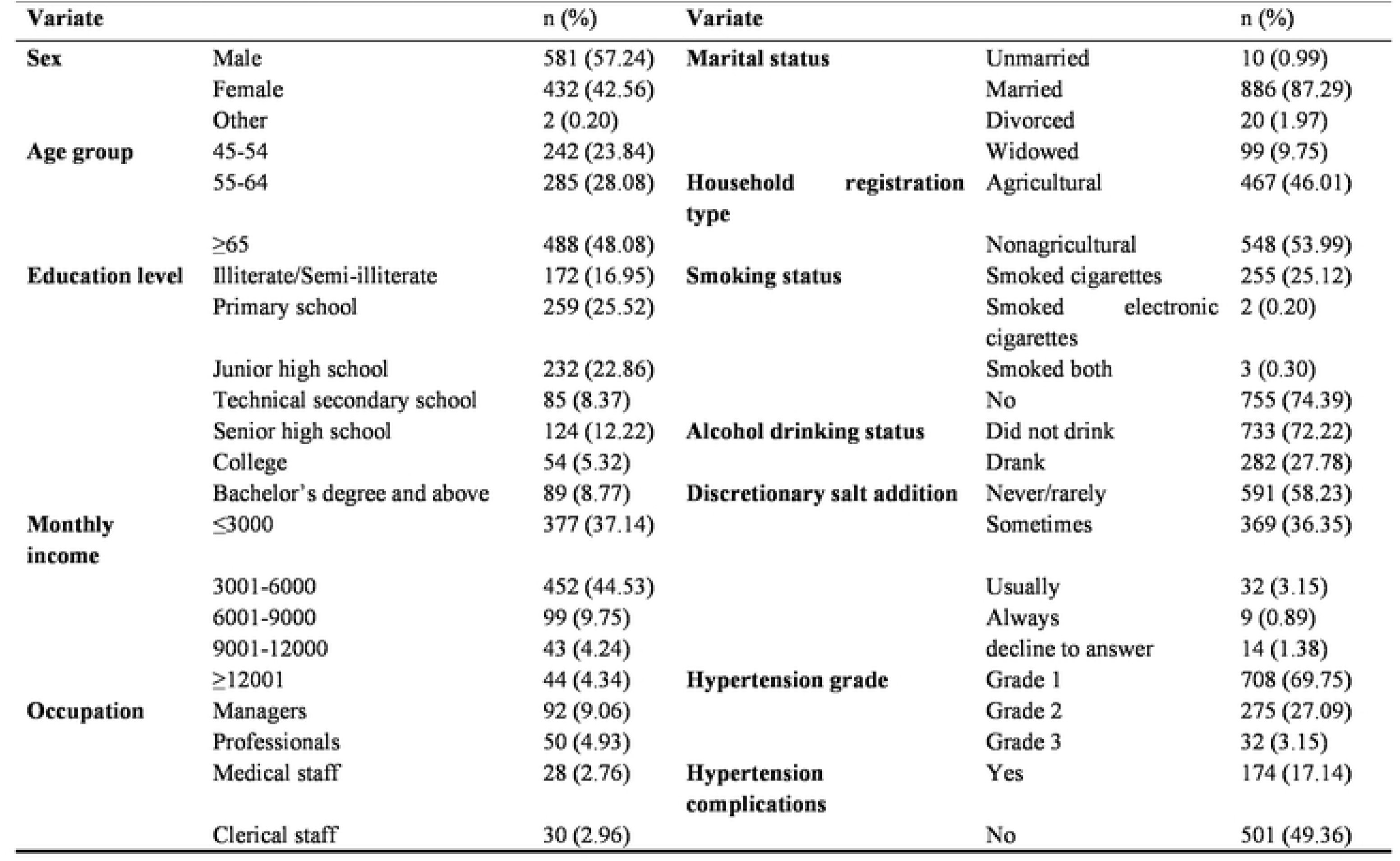
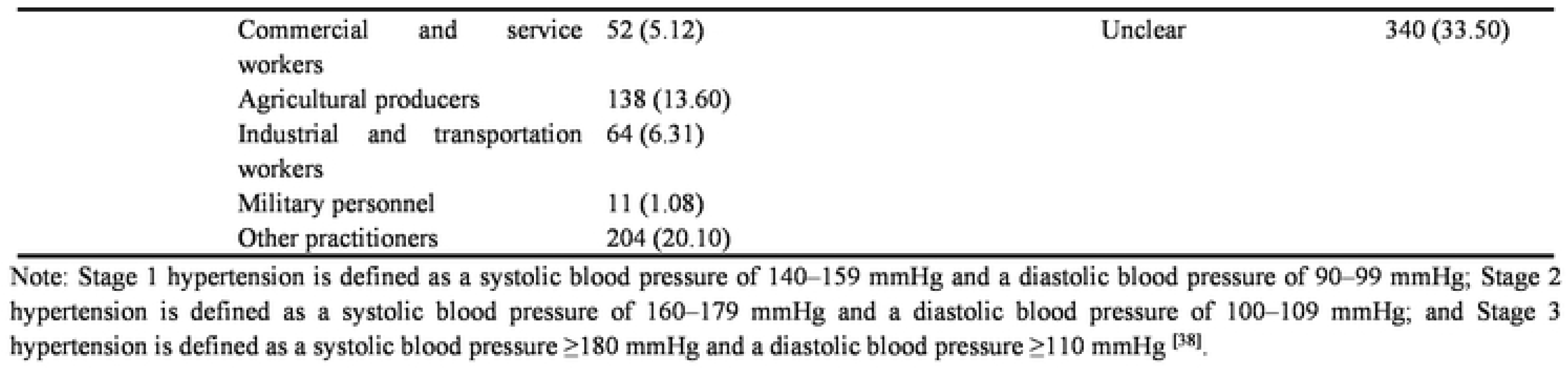
Basic characteristics of the respondents.

### Perceived social support, self-efficacy, health literacy, and quality of life scores

The total score for perceived social support was 15.57 ± 3.45, with the dimension “receiving emotional support from family when needed” scoring the highest (5.38 ± 1.24), while the dimension “having people in life who care about my feelings” scored slightly lower (5.02 ± 1.39). The total self-efficacy score was 10.61 ± 2.41, with scores across all dimensions ranging from 3.48 to 3.61. The overall health literacy score was 9.49 ± 2.86, with the dimension “accessing disease treatment information” scoring the lowest (2.49 ± 0.80). Regarding quality of life, the EQ-5D utility index score (0.88 ± 0.18) was relatively low across all dimensions (1.22 to –1.63), with only the “pain or discomfort” dimension scoring significantly higher (1.63 ± 0.63). The EQ-VAS score (71.06 ± 17.49) fell within the upper-middle range.

### Correlation Analysis Between Perceived Social Support, Self-Efficacy, Health Literacy, and Quality of Life

The correlation analysis results demonstrated that perceived social support, self-efficacy, health literacy, and the EQ-5D utility index showed positive correlations with the EQ-VAS variables. Specifically, perceived social support exhibited a weak but statistically significant positive correlation with EQ-VAS (r=0.17, p<0.001); self-efficacy showed a low but statistically significant positive correlation with EQ-VAS (r=0.28, p<0.001); health literacy demonstrated a low but statistically significant positive correlation with EQ-VAS (r=0.24, p<0.001); and the EQ-5D exhibited a moderately strong positive correlation with EQ-VAS (r=0.42, p<0.001) (Figure 1).

**Figure 1.**
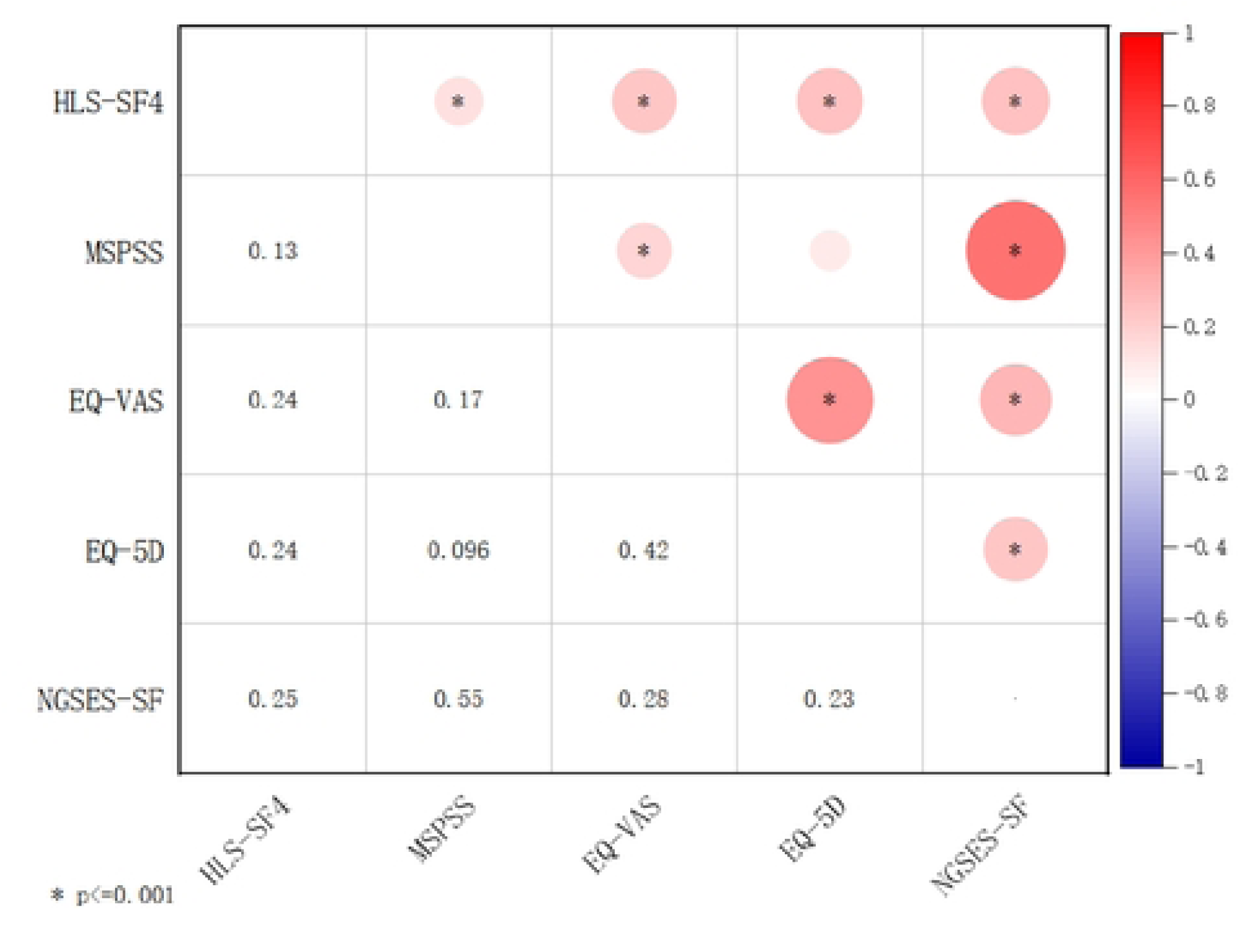
Correlation Heatmap of Key Study Variables.

### The Chain-Mediating Effect of Perceived Social Support and Self-Efficacy on the Relationship Between Health Literacy and Quality of Life

This study employed health literacy as the independent variable, quality of life (measured by the EQ-5D utility index and EQ-VAS) as a dependent variable, and self-efficacy and perceived social support as mediating variables. The analysis was conducted using a chain mediation structural equation model, with model optimization and fitting performed via maximum likelihood estimation to validate hypotheses. The fit indices for the revised model were as follows: χ²/df = 7.773, RMSEA = 0.082, GFI = 0.933, TLI = 0.916, and SRMR = 0.050. These values indicate that the model is acceptable. The model structure and standardized path coefficients are presented in Figure 2. In this model, health literacy, self-efficacy, and perceived social support were defined as latent variables (each measured through 3–4 observed indicators). Regarding quality of life, the EQ-5D utility index comprised five observed indicators, whereas EQ-VAS used a single observed indicator. Path coefficient analysis revealed that health literacy had a positive direct effect on both self-efficacy (path coefficient, 0.591) and perceived social support (path coefficient, 0.134). Additionally, self-efficacy positively influenced EQ-VAS scores (path coefficient, 0.239), while perceived social support was positively correlated with self-efficacy (path coefficient, 0.907). Furthermore, health literacy indirectly influenced quality of life through the mediating variables.

**Figure 2.**
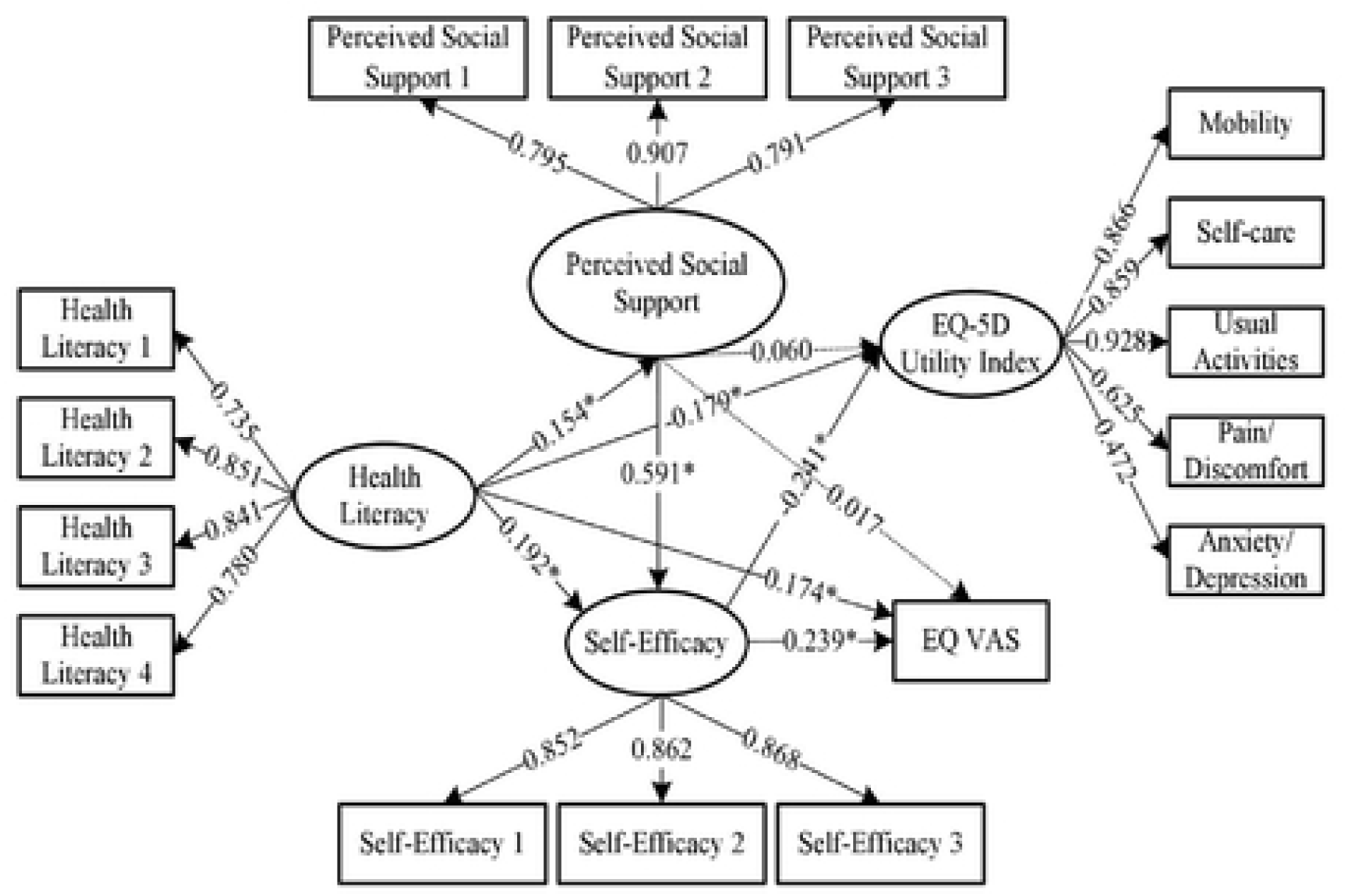
Structural equation model. Standardized path coefficients are shown. (*p < 0,001)

The mediation model was tested using the self-service method (5,000 repeated samplings). The results showed that the 95% confidence intervals for all pathways did not include zero (p <0.001), indicating statistically significant mediation effects. Regarding the impact on the EQ-5D utility index, the total effect size was-0.238 (the 95% confidence interval did not include zero, indicating a significant effect), with the direct effect (-0.179) accounting for 75.21% of the total effect. In the mediation analysis, only the pathway “Health Literacy → Self-Efficacy → EQ-5D Utility Index” (effect size: -0.046, accounting for 19.33%) and the pathway “Health Literacy → Perceived Social Support → Self-Efficacy → EQ-5D Utility Index” (effect size: -0.022, accounting for 9.24%) were statistically significant. The pathway “Health Literacy → Perceived Social Support → EQ-5D Utility Index” was not significant (the 95% confidence interval included zero). The total effect size for EQ-VAS was 0.245 (statistically significant), with the direct effect (0.174) accounting for 71.02% of the total effect. In terms of mediation effects, both the pathway “Health Literacy → Self-Efficacy → EQ-VAS” (effect size: 0.046, accounting for 18.78% of the total effect) and the pathway “Health Literacy → Perceived Social Support → Self-Efficacy → EQ-VAS” (effect size: 0.022, accounting for 8.98% of the total effect) were significant. The pathway “Health Literacy → Perceived Social Support → EQ-VAS” did not reach statistical significance. The results are presented in Table 2.

**Table 2.** Results of path analysis.

| Path relationship | Effect size | SE | 95% CI | Effect proportion |
| --- | --- | --- | --- | --- |
| Overall effect (Health Literacy → EQ-5D Utility index) | -0.238 | 0.041 | -0.313~-0.155 | |
| Direct effect (Health Literacy → EQ-5D Utility index) | -0.179 | 0.042 | -0.258~-0.095 | 75.21% |
| Health Literacy → Perceived Social Support → EQ-5D Utility Index | 0.009 | 0.009 | -0.006~0.031 | |
| Health Literacy → Self-Efficacy → EQ-5D Utility Index | -0.046 | 0.015 | -0.081~-0.023 | 19.33% |
| Health literacy → Perceived Social Support → Self-efficacy → EQ-5D Utility Index | -0.022 | 0.008 | -0.041~-0.010 | 9.24% |
| Overall effect (Health Literacy → EQ-VAS) | 0.245 | 0.036 | 0.173~0.313 | |
| Direct effect (Health Literacy → EQ-VAS) | 0.174 | 0.038 | 0.100~0.247 | 71.02% |
| Health Literacy → Perceived Social Support → EQ-VAS | 0.003 | 0.007 | -0.011~0.018 | |
| Health Literacy → Self-Efficacy → EQ-VAS | 0.046 | 0.013 | 0.024~0.077 | 18.78% |
| Health Literacy → Perceived Social Support → Self-Efficacy → EQ-VAS | 0.022 | 0.008 | 0.010~0.040 | 8.98% |

## Discussion

In this study, the patients’ quality of life scores were 0.88±0.18 (EQ-5D utility index) and 71.06±17.49 (EQ-VAS), indicating a moderate quality of life. These scores were lower than those reported by Wang et al. ^[31]^. This discrepancy may arise from the age of the hypertensive patients in this study, who were middle-aged and older, for whom hypertension constitutes a significant adverse event. Under the dual impacts of aging and this negative health condition, quality of life in this population undergoes changes accordingly. Middle-aged and older hypertensive patients with higher health literacy demonstrated enhanced cognitive awareness and utilization of social support, thereby strengthening their self-efficacy. This increase in self-efficacy, in turn, motivated these individuals to proactively adopt a healthier lifestyle, creating a positive cycle that improved their overall quality of life.

Hypertension is often referred to as the “silent killer.” In this study, 69.75% of stage 1 hypertensive patients exhibited no obvious symptoms and no significant abnormalities in quality of life, while 50.64% of middle-aged and older hypertensive patients had hypertension-related complications or provided unclear responses regarding their complication status. The EQ-5D utility index and its dimension scores indicated that core dimensions such as mobility and self-care capacity were in the low-utility range, with only the pain/discomfort dimension scoring relatively high. In this study, 17.14% of patients developed hypertension-related complications, and the occurrence of these complications was associated with accelerated functional decline ^[32]^. Patients with hypertension-related complications scored significantly lower than those without these complications with regard to the dimension of mobility, pain/discomfort, and activities of daily living dimensions of the EQ-5D-5L scale.

Perceived social support is positively correlated with quality of life in hypertensive patients. The dimension “obtaining familial emotional support when needed” scored the highest, whereas the dimension “feeling care from others in daily life” exhibited a lower score. This finding confirms that family is the primary source of perceived social support for hypertensive individuals, and emotional care from friends and other social relationships can be enhanced through strengthened emotional bonds. Enhancing the provision of emotional care from friends and other social relationships can better leverage the promoting effect of perceived social support on the health of hypertensive patients.

Cross-sectional studies on hypertensive patients have demonstrated a significant correlation between health literacy levels and quality of life ^[33,34]^. Middle-aged and older individuals are particularly susceptible to insufficient health literacy due to limited education, low self-care awareness, an inadequate understanding of modern healthcare, and the influence of traditional beliefs. The interplay between their health status and poor health literacy exacerbates the challenges in managing hypertension, increases the risk of complications, and decreases quality of life among this demographic. In this study, the average overall health literacy score among patients was merely 9.49 ± 2.86, with the lowest score recorded for the “access to disease treatment information” dimension (2.49 ± 0.80). Insufficient health information access is associated with impaired self-management capabilities and unhealthy behaviours and thereby adversely affects quality of life ^[35]^.

Health literacy and quality of life are related via mediating mechanisms in which perceived social support and self-efficacy exhibit distinct mediating effects. This study reveals that self-efficacy significantly mediates the relationship between health literacy and quality of life, accounting for 19.33% of the total effect on the EQ-5D utility index and 18.78% of the total effect on the EQ-VAS score. A study conducted among the Chinese population also confirmed similar findings ^[36]^. This mediating relationship between health literacy and quality of life may be attributable to the idea that patients with higher health literacy demonstrate stronger self-discipline and adopt more proactive self-health management strategies. Such patients are better able to understand and apply health guidance, engage fully in self-care, exhibit higher self-efficacy, and ultimately achieve significant improvements in their quality of life ^[37]^. The mediating path of perceived social support between health literacy and quality of life was not statistically significant, indicating that this path does not independently mediate this relationship. Instead, the association between social support and quality of life may be mediated by variables such as self-efficacy.

## Limitations

This study has several limitations. First, the survey participants were selected from six central provinces of China—Shanxi, Anhui, Jiangxi, Henan, Hubei, and Hunan Provinces—which may have introduced selective bias. Second, although this cross-sectional study only analysed the mediating effects of perceived social support and self-efficacy on health literacy and quality of life among middle-aged and older hypertensive patients, a rigorous mixed-methods research design combining cross-sectional surveys and qualitative interviews is required to further elucidate the mechanisms underlying this relationship. Third, the study did not incorporate comorbidity factors among hypertensive patients; comorbidities may influence the correlations between research variables, thereby limiting the interpretability of the findings. Fourth, multiple psychosocial constructs were measured using brief scales. Although the reliability and validity of these scales have been validated, they may still exhibit limitations such as insufficient construct depth and representativeness, which could affect the interpretation and generalizability of the results. Fifth, the structural equation model reported in this study focused solely on the path relationships among four core latent variables (health literacy, perceived social support, self-efficacy, and quality of life); future research could include additional covariates.

## Conclusion

Health literacy is significantly correlated with quality of life in middle-aged and older hypertensive patients and plays a crucial explanatory role in improving quality of life through a chain-mediated pathway involving perceived social support and self-efficacy. For these patients, both perceived social support and self-efficacy levels show a significant positive correlation with improved quality of life. The synergistic enhancement of social support and self-efficacy is positively correlated with the adoption of an active and healthy lifestyle among middle-aged and older individuals, thereby substantially promoting overall quality of life.

## Data Availability

The raw data has been uploaded to the system in Appendix 1 format.

## Acknowledgement

We thank American Journal Experts for English language editing.

## Contribution

All author contributed equal.n

## Human Ethics and Consent to Participate declaration

Not applicable.

## Consent to Participate declaration

All participants signed the Consent Forms.

